# Toward Automated Classification of Pathological Transcranial Doppler Waveform Morphology via Spectral Clustering

**DOI:** 10.1101/19003236

**Authors:** Samuel G. Thorpe, Corey M. Thibeault, Nicolas Canac, Kian Jalaleddini, Amber Dorn, Seth J. Wilk, Thomas Devlin, Fabien Scalzo, Robert B. Hamilton

## Abstract

Cerebral Blood Flow Velocity waveforms acquired via Transcranial Doppler (TCD) can provide evidence for cerebrovascular occlusion and stenosis. Thrombolysis in Brain Ischemia (TIBI) flow grades are widely used for this purpose, but require subjective assessment by expert evaluators to be reliable. In this work we seek to determine whether TCD morphology can be objectively assessed using an unsupervised machine learning approach to waveform categorization. TCD beat waveforms were recorded at multiple depths from the Middle Cerebral Arteries of 106 subjects; 33 with CTA-confirmed Large Vessel Occlusion (LVO). From each waveform, three morphological variables were extracted, quantifying absolute peak onset, number/prominence of auxiliary peaks, and systolic canopy length. Spectral clustering identified groups implicit in the resultant three-dimensional feature space, with gap-statistic criteria establishing the optimal cluster number. We found that gap-statistic disparity was maximized at four clusters, referred to as flow types I, II, III, and IV. Types I and II were primarily composed of control subject waveforms, whereas types III and IV derived mainly from LVO patients. Cluster morphologies for types I and IV aligned clearly with Normal and Blunted TIBI flows, respectively. Types II and III represented commonly observed flow-types not delineated by TIBI, which nonetheless deviate quantifiably from normal and blunted flows. We conclude that important morphological variability exists beyond that currently quantified by TIBI in populations experiencing or at-risk for acute ischemic stroke, and posit that the observed flow-types provide the foundation for objective methods of real-time automated flow type classification.

## I. Introduction

Transcranial Doppler ultrasound (TCD) is a noninvasive methodology for measuring Cerebral Blood Flow Velocity (CBFV) through the large arteries of the brain. The morphology of the pulsatile CBFV waveform can provide information concerning numerous cerebrovascular pathologies [1], including stroke [2–4], intracranial hypertension [5], and mild Traumatic Brain injury [6]. TCD has proven effective for detecting occluded and stenosed cerebral arteries in the context of acute ischemic stroke [7–9]. In these studies, stroke pathology is primarily quantified using Thrombolysis in Brain Ischemia (TIBI) flow grades to evaluate waveform morphology [10], with waveform features such as onset of maximum velocity, and evidence of standard pulsatile and peak structure evaluated for grade assignment. Numerous such studies have shown TIBI assessment to be a valuable tool for these purposes, possessing sensitivity and specificity often exceeding 90%. [2,9,11] However, reliable determination of TIBI grades requires subjective assessment by experts, severely limiting their utility for prehospital stroke assessment by less specialized personnel. Recent work by our group has shown that a TCD-derived morphological biomarker termed Velocity Curvature Index (VCI) may provide a robust, objectively computable metric for detecting Large Vessel Occlusion (LVO) [12,13]. Though VCI readily identifies waveforms with pathologically deviant curvature, it does not differentiate between pathological morphologies such as those delineated by the TIBI scale. An objective means of waveform categorization could thus provide additional information concerning stroke etiology to better inform stroke triage and transfer decisions.

The TIBI categories range from grades 0 - 5, with 5 indicating normal flow, and grades 0 and 1 designating absent (0) or minimal (1) flow associated with complete or partial vascular occlusion. The grades between are associated with characteristic morphologies which are used to determine blunting (grade 2), dampening (grade 3), or stenotic (grade 4) flows. The complexity and subjectivity in assignment of these categories, along with their evident utility for stroke assessment, make them a natural candidate problem for automation via machine learning. However, before attempting this explicitly, we feel a more foundational problem must first be addressed, in that it is currently unclear to what degree these categories actually capture the natural variance inherent in empirical data. That is, we know these categories are useful, but we do not know that they are comprehensive. Additional waveforms may be present which are also informative, or perhaps a different subset may better explain variability across subjects.

In this work we take a data-driven approach to waveform categorization, retrospectively applying an unsupervised learning algorithm to a dataset comprised of multiple subject groups, including patients experiencing acute LVO, as well as control subjects collected both in and out of hospital. For these purposes we employ Spectral Clustering [14,15], which does not make strong assumptions about inherent cluster density, and thus performs well when clusters are connected but potentially non-convex. We hypothesize that LVO subject waveforms will fall into clusters which are mostly distinct from non-LVO controls, for which we can subsequently compare alignment with established TIBI categories.

## II. Materials and Methods

### A. Subjects

We compared TCD waveform morphology across three subject groups; one with CTA-confirmed LVO, a second non-LVO control group collected in-hospital, and a third group of control subjects collected out of hospital. LVO and in-hospital controls (IHC) were enrolled at Erlanger Southeast Regional Stroke Center in Chattanooga, TN, between October 2016 and October 2017. Clinical and imaging methodological details for these subjects are given in [12,13]. Out of hospital controls (OHC) were scanned at multiple locations in Los Angeles, CA. Volunteers were eligible for enrollment if between 40 – 85 years of age, and no exclusion criteria applied (Table 1). Experiment protocols for LVO and IHC subjects were approved by University of Tennessee College of Medicine Institutional Review Board (ID: 16-097), and for OHC subjects by Western IRB (ID: 20151682).

**TABLE 1.** Subject Exclusion Criteria.

| LVO, IHC Subjects | OHC Subjects |
| --- | --- |
| 1) Head CT findings consistent with acute primary intracranial hemorrhage (SAH, ICH, etc.). | 1) Individuals taking any psychoactive medication. |
| 2) Patients requiring pharmacological support for hypotension. | 2) Systolic arterial blood pressure greater than 140 mmHg |
| 3) Subjects who underwent partial or full craniotomy. | 3) Individuals presenting with an external wound in head. |
| 4) Additional intracranial pathologies present (tumor, hydrocephalus, etc.). | 4) Pregnant Women. |
| 5) Anticipated insufficient time to acquire a complete set of scan as described by the protocol. | 5) Individuals who have a known history of: severe TBI, moderate TBI (within 3 years), vascular disease, stroke, sickle cell anemia, brain tumor, epidural or subdural hematoma, abnormal MRI or CT scan, cardiovascular disease. |
| 6) Significant hemodynamic pharmacological agent (cocaine, amphetamine, etc.). |  |
| 7) Subjects who are under arrest for a felony. |  |

Subject exclusion criteria. Note the following abbreviations: SAH – Subarachnoid Hemorrhage, ICH – Intracranial Hemorrhage, TBI – Traumatic Brain Injury, MRI – Magnetic Resonance Imaging, CT – Computerized Tomography.

### B. TCD Waveform Recording & Processing

CBFV signals were acquired by trained technicians using 2 MHz handheld probes to transtemporally insonate the left/right MCA. The technician was instructed to obtain recordings for as many depths as possible between 45 – 60 mm in both the left/right cerebral hemispheres. Once signal was identified and optimized at a specific depth, waveform recordings were made in 30-second intervals. Individual beat waveforms from each recorded depth were extracted offline (post-recording) using an automated beat identification algorithm [16], with automated beat outlier rejection via Iterated Interquartile Range exclusion with cross-correlation and beat length as primary comparators [17]. To be included in this analysis, exams were required to contain at least one bilateral pair of left/right MCA scans at depths between 45 – 60 mm, each containing at least 15 accepted beats. Accepted beats were aligned and averaged, resulting in a single representative beat waveform for each recorded interval. OHC waveforms, digitally sampled at 400 Hz, were resampled to 125 Hz to match the native sampling rate of LVO and IHC waveforms. All waveforms were smoothed via convolution with a 9 ms Hanning window to reduce high-frequency noise. Since this analysis sought to evaluate morphological commonality regardless of underlying heart rate or velocity scale, each waveform was normalized with respect to both time and velocity. The velocity normalization was accomplished for each waveform by first subtracting the minimum, then subsequently dividing by the resultant maximum (thus rescaling to the interval [0, 1] along the velocity axis). The temporal normalization was accomplished The temporal normalization was accomplished by resampling each waveform to 100 total samples (via cubic spline), effectively enforcing a common heart rate across waveforms.

### C. Cluster Feature Extraction

From each waveform, denoted x(t) in equations 1 – 3, we extracted three morphological features (figure 1). The first, onset (equation 1), marked the temporal onset of maximal velocity. Canopy (equation 2), was defined as the number of samples comprising the beat “canopy”, i.e. the cardinality (denoted *card*) of the set of samples with velocity greater than 25% of the diastolic-systolic range [see 12 for details]. The final feature, peaks (equation 3), quantified the number and “weight” of waveform peaks. First we identified the set of true-peaks (TP, approximate zeros of the first derivative) as points in the canopy corresponding to a sign-change in the difference between successive samples. True peaks were each assigned a weight of one. Next we identified the set of “pseudo-peaks” (PP, points where the derivative is small but non-vanishing) where the difference magnitude between successive samples dropped below a critical threshold of 0.01 (choosing the point with smallest difference magnitude in any group of adjacent sub-threshold samples). For pseudo-peaks we assigned weights corresponding to one minus the ratio of the associated difference magnitude and the threshold 0.01, such that those with the smallest derivative were weighted most heavily. The peaks variable was then computed as the sum over all corresponding weights.

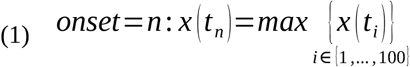

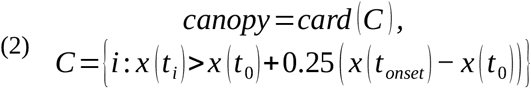

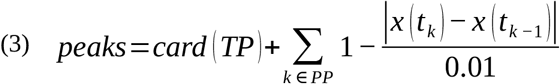

**Fig. 1.**
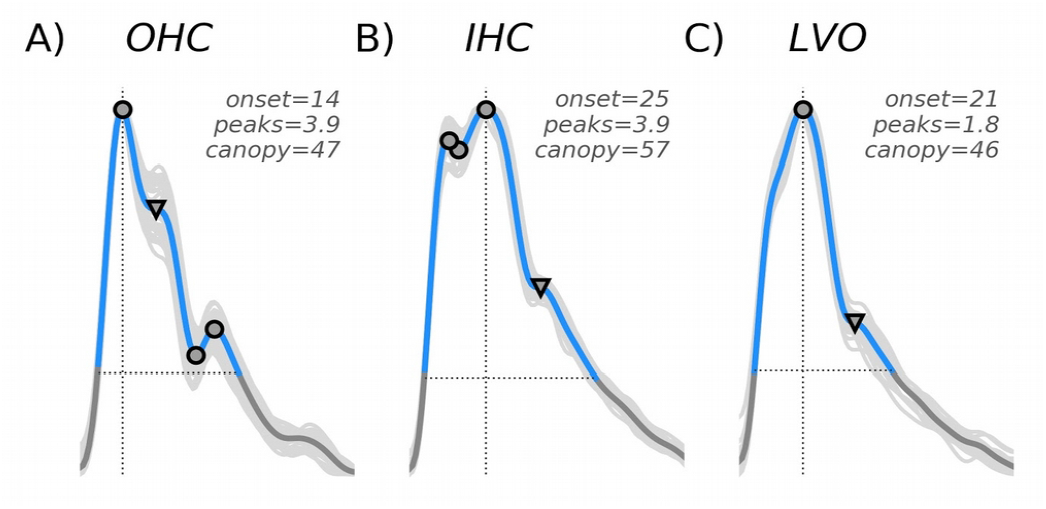
Cluster variables are depicted for three example waveforms taken from each of the subject groups; OHC (A), IHC (B), and LVO (C). All waveforms were normalized in both time and velocity, so as to span 100 total samples ranging from zero to one on the y-axis. The onset variable (vertical line) marks the time sample where maximum velocity is attained. The canopy variable (horizontal line) marks the length (in samples) of the systolic canopy. The peaks variable is a weighted sum of waveform peaks, both true (indicated by circles) and pseudo (triangles).

### D. Spectral Clustering and Gap Analysis

The extracted features were z-transformed, and the resultant three-dimensional feature space was partitioned via spectral clustering implemented in Scikit-learn [18], specifically the SpectralClustering module with default parameters and radial basis kernel. To determine the optimal number of clusters, gap statistics (Gk) were computed for total clusters (k) ranging from two to seven. In this procedure, gap statistics are computed as the difference between observed log intra-cluster dispersion pooled across k clusters (denoted Wk), and the analogous expected dispersion bootstrapped from a null distribution incorporating the covariance structure of the observed data [19]. Each of the 1000 bootstrap iterations was generated by sampling uniformly over the range of the columns of the observed data transformed by its right-singular vectors, and back-transforming the resultant sample to feature space via the right-singular transpose (method b in [19]). The optimal number of clusters were selected as the smallest k such that Gk > Gk+1 – Sk+1, where Sk is the standard deviation of the k-cluster bootstrap distribution corrected to account for simulation error.

### E. Cluster Archetypes

To visualize the characteristic morphology of the resultant clusters, archetypal waveforms were derived for each by computing the matrix of correlation distances between all cluster member waveforms, and ranking by average distance to other members. The five waveforms with smallest mean intra-cluster distance were averaged to obtain the waveform archetype for each cluster.

## III. Results

### A. Subject Demographics

The current analyses included 33 LVO subjects (16 female), 33 IHC subjects (13 female), and 40 (25 female) OHC subjects, with average ages of 66.9 (SD=15.7), 56.4 (SD=16.3), and 58.4 (SD=10.9) years, respectively. A total of 996 average beat waveforms were included in this analysis, with 354, 445, and 196 contributed by LVO, IHC, and OHC subjects, respectively.

### B. Gap Analysis and Cluster Archetypes

Figure 2A shows expected and observed log intra-cluster dispersion as a function of total clusters, with the gap statistic determined by their difference. The elbow in observed dispersion at four clusters corresponds to both the maximum gap statistic (figure 2B) and optimal number of clusters determined by the Tibshirani et al. (2001) criteria. The associated feature space (figure 3A) is shown partitioned into the four resultant clusters, with membership indicated by color. The largest cluster (type I), containing 400 waveforms, was characterized by early max velocity onset with wide canopy and strong peaks (figure 3B). Of waveforms in this cluster, 18% came from LVO subjects, with 82% from controls (52% IHC, 30% OHC). The second largest cluster (type II), containing 257 waveforms, was characterized by later max velocity onset, with wide canopy and strong peaks (figure 3C). Of waveforms in this cluster, 20% came from LVO subjects versus 80% from controls (68% IHC, 12% OHC). The smallest cluster (type III), containing 83 waveforms, was characterized by early max velocity onset, with narrow canopy and weak peaks (figure 3D). Of waveforms in this cluster, 95% came from LVO subjects versus 5% from controls (1% IHC, 4% OHC). The final cluster (type IV), containing 256 waveforms, was characterized by late max velocity onset, with wide canopy but weak peaks (figure 3E). Of waveforms in this cluster, 60% came from LVO subjects versus 40% from controls (28% IHC, 12% OHC).

**Fig. 2.**
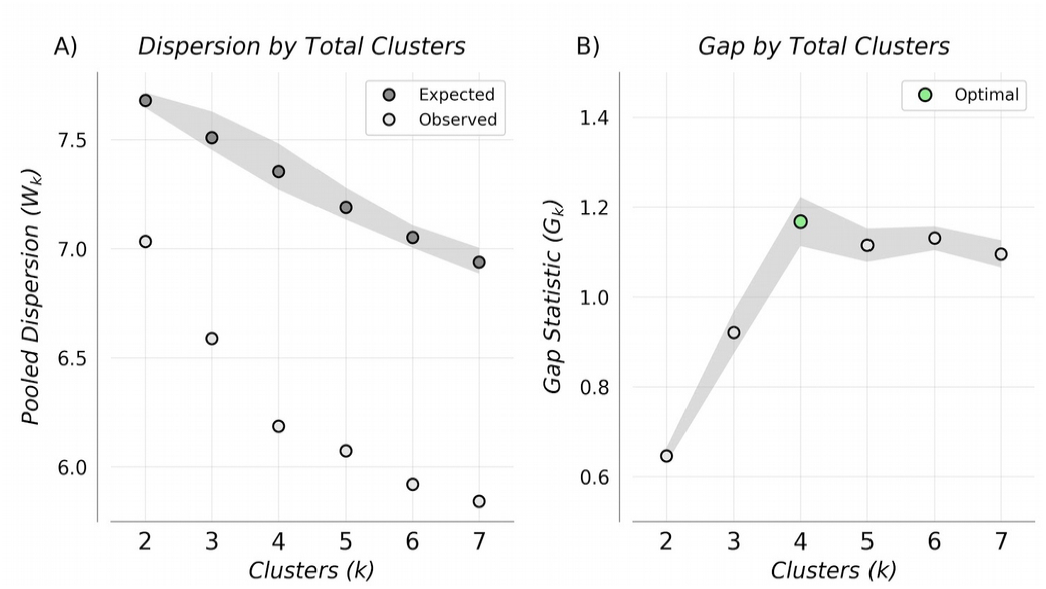
Pooled Intra-cluster dispersion (A), and associated Gap Statistics (B) as a function of cluster number. Gap-statistic disparity was maximized at four clusters which also corresponded to the optimal number given by the selection criteria in Tibshirani et al. [19]

**Fig. 3.**
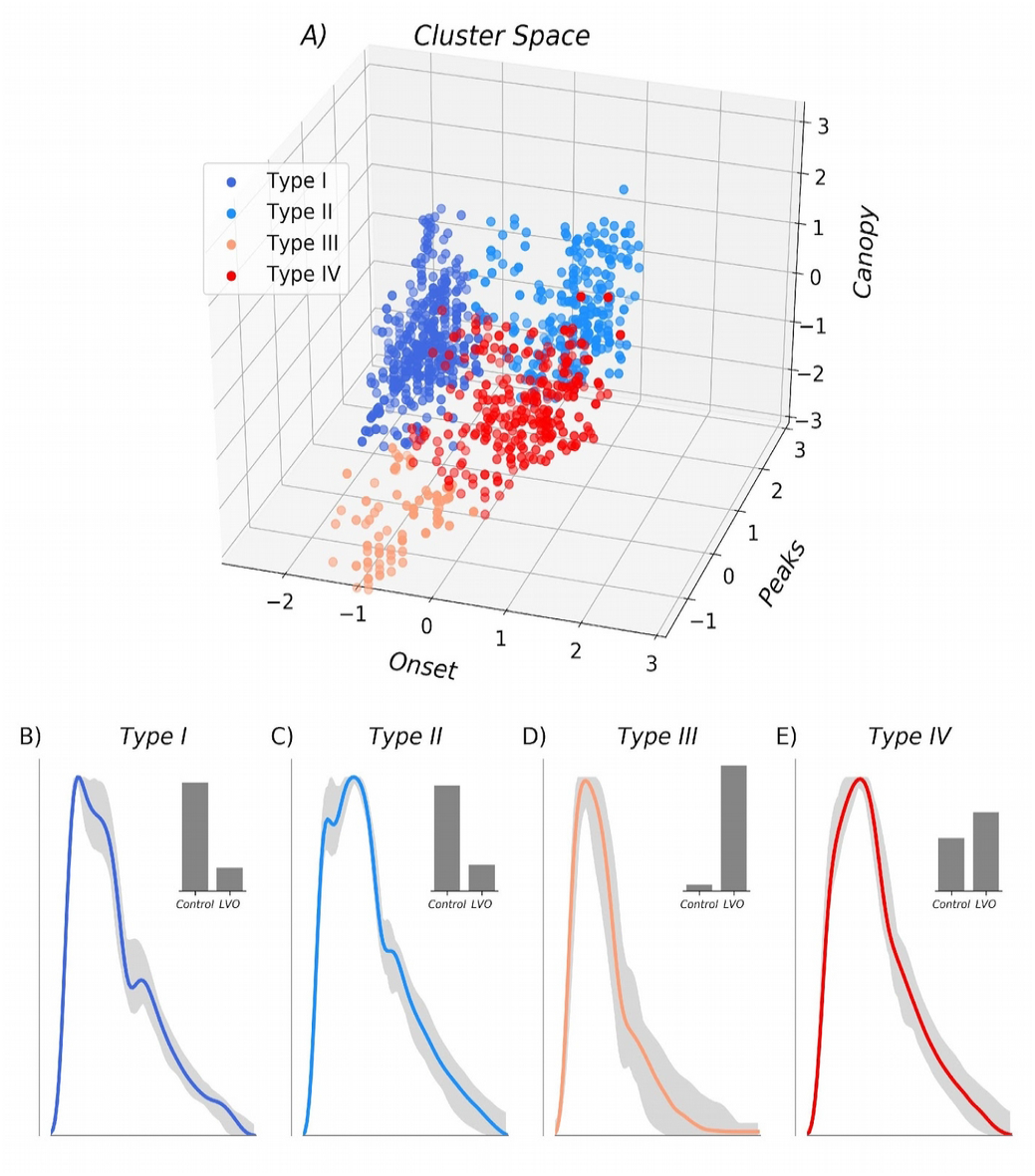
Three-dimensional feature space is shown in Z-scored coordinates with the optimal four clusters indicated by color. Associated cluster morphologies are shown for each beat type (B-E), with the most representative exemplar for each cluster shown in color, and the range of the next 50 most representative variants depicted in gray. Associated histograms in the upper right (B-E) demonstrate that Type I and II clusters were primarily composed of waveforms from control patient populations, whereas Type III and IV clusters were primarily composed of LVO patient waveforms.

**Fig. 4.**
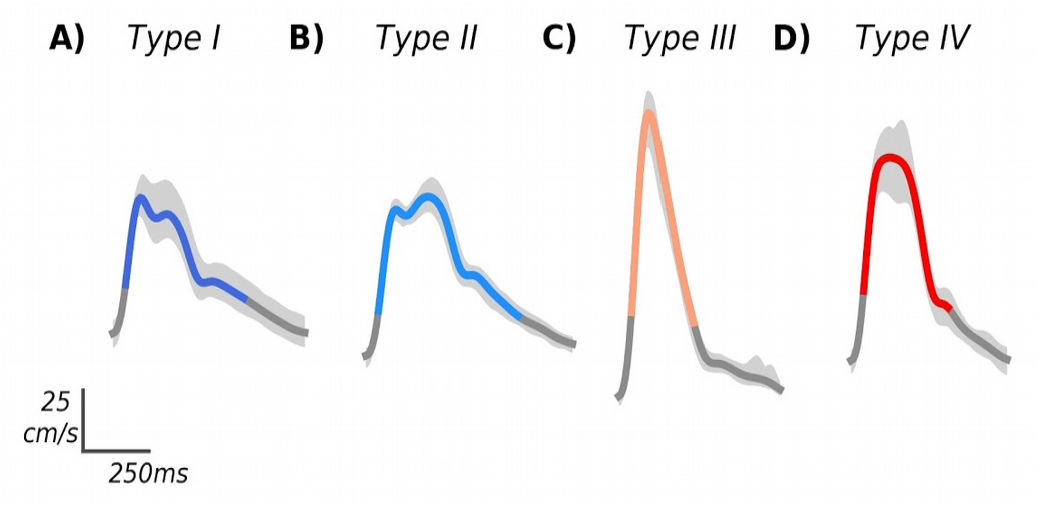
Individual example waveforms from each cluster, shown in standard units (unnormalized), with variability across individual beats depicted in light gray. The type I and II examples (A and B) were sampled from the IHC group, whereas the types III and IV examples (C and D) were selected from the LVO subjects.

## IV. Discussion

### A. Comparison to TIBI Flow Grades

Of the clusters we observed, two have definitive analogues on the TIBI scale. Specifically, our type I cluster showed the early systolic maximum and recognizable peak structure associated with TIBI grade 5 normal flow. Similarly, our type IV cluster exhibited the delayed flow acceleration with no discernable early peak, and maximum velocity in mid-to-late systole, characteristic of TIBI grade 2 blunted flow. Accordingly, we observed type I flows more often in control subjects, whereas type IV flows were more commonly associated with LVO. The remaining clusters did not have unambiguous TIBI analogues, though their contrasting subject group compositions suggest type II is more commonly observed in controls, and type III nearly always associated with LVO. The type II cluster, characterized by late onset maximal velocity but otherwise normal peaks, may reflect differences in peripheral vascular resistance relative to type I, which could conceivably impact either or both the initial systolic upstroke and/or the timing of reflected waves affecting the amplitude of the mid-systolic peak.

Interpretation of the pathological type III morphology represents the most speculative aspect of this work. We infer from its strong association with LVO subjects that it likely results from occlusion or stenosis of the cerebral vessels, though in a manner distinct from typically blunted waveforms, leaving the initial systolic acceleration unaffected while suppressing all subsequent morphological structure; features which are especially clear in the unnormalized subject waveforms (see figure 4 for examples). This pattern is intriguingly similar to previously observed waveforms associated with vascular stenosis (see [20] figure 4c), suggesting that this flow type may rightly be thought of as the analog of the TIBI stenotic grade 4. However, multiple reasons compel us to be cautious with this claim. First, this flow type was by far the smallest cluster, which manifest in only a small number subjects. Case reports for most of these subjects did indicate significant, but non flow-limiting stenoses. However, such moderate stenoses are not uncommon in the LVO subject population in general. Moreover, the TIBI grade 4 categorical definition given by Demchuk et al. [10] refers both to elevated velocity relative to the adjacent (unaffected) hemisphere, which are not clearly born out in our subjects, as well as evidence for flow turbulence, which spectrograms are required to assess. For the time being we reserve judgement as to whether future studies incorporating richer spectral data might confirm this flow type to have stenotic etiology.

### B. Limitations and Future Work

Considering the remaining TIBI flow grades, the lowest are not associated with sufficiently pulsatile CBFV waveforms, and thus could not be represented in our data set. Specifically, grade zero is defined as the absence of flow, whereas grade 1 (minimal flow) is so weakly pulsatile as to give rise to nearly flat waveforms when averaged over successive beats. In this study we required subjects to exhibit bilateral pulsatile data to be included in analysis. This was done to ensure that absence of temporal acoustic windows would not be mistaken for absence of MCA flow. However, future experimental protocols could be modified to require TCD examination of all cerebral vessels in the anterior circulation. Evidence of flow in any of these vessels in a given hemisphere without corresponding evidence of adjacent MCA flow would allow for confident assessment of absent or minimal flow grades, likely bringing our current results further into alignment with the TIBI scale.

The remaining TIBI grade 3 flow (Dampened), is not solely morphologically defined, requiring comparison of velocity magnitude relative to a control waveform for assignment, and thus cannot clearly align with our clusters. Future work could explore whether our clustering framework might be extended for application to sets of waveforms, including relative velocities as features, which might help reconstruct these other TIBI categories.

From the 3-dimensional cluster space (figure 3A) it is clear that adjacent cluster types II and IV share a fuzzy boundary primarily determined by the peaks variable, suggesting the two can be difficult to differentiate when systolic peaks are not clear. Indeed, the type IV cluster had the least homogenous group composition, with 40% of waveforms originating from control subjects; a fact which would negatively impact specificity were we to use these clusters alone to classify LVO. Clearly, further work is needed to determine whether additional or refined clustering features, perhaps derived from waveform spectrograms and associated Mmode, might help disambiguate these groups.

## V. Conclusions

To our knowledge, this study presents the first unsupervised learning analysis of LVO pathology evident in the TCD signal. We have shown that spectral clustering can readily recover meaningful TCD flow types bearing clear relation to known morphological categories. Moreover, the resultant cluster archetypes provide definitive morphological templates, enabling automated categorization of novel waveforms via numerous potential comparative methods, the refinement of which should provide fruitful avenues for future work. Ultimately, we will explore whether such automated labels can be combined with other established metrics, such as VCI and velocity asymmetry, to improve LVO classification efficacy.

## Data Availability

Data used for all visualizations and statistical comparisons within the manuscript are available on Figshare data repository at:
https://figshare.com/articles/TCD_Clustering_Data/9037388
Additional materials may be made available by specific request to the corresponding author.

